# Choroid plexus calcification detection using quantitative susceptibility mapping MRI

**DOI:** 10.64898/2026.05.26.26354154

**Authors:** Kilian Hett, Abigail Dubois, Isabelle Bonitz, Ciaran M. Considine, James Eaton, Colin D. McKnight, Daniel O. Claassen, Manus J. Donahue, Paula Trujillo

## Abstract

**Purpose:** The choroid plexus (ChP) is the primary source of cerebrospinal fluid and an emerging marker of cerebral health, with enlargement and hypoperfusion reported in aging and neurodegeneration. However, frequent ChP calcifications can confound volumetric and perfusion measures. Although computed tomography (CT) is the gold standard for detecting calcification, it is rarely available in research MRI. Quantitative susceptibility mapping (QSM) offers an alternative sensitive to diamagnetic mineralization but lacks validated susceptibility thresholds.

**Method:** Participants underwent CT and MRI within four weeks, including 3D T_1_-weighted and a multi-echo gradient echo QSM MRI. ChP calcifications were identified on CT using standard diagnostic criteria. Using the Bayes decision boundary framework, we identified optimal susceptibility thresholds for detecting diamagnetic signals consistent with calcification and compared these thresholds with multiple density levels measured on gold standard CT images.

**Results:** Across all participants (n=20; age=62.2±12.0 yrs), the optimal susceptibility threshold separating background ChP signal from calcifications was -0.10 ppm at 60 HU (low-density) and -0.15 ppm at 100 HU (high-density). Susceptibility values within calcified tissue exhibited a linear relationship with CT-derived tissue density. A significant positive association was observed between ChP volume and calcification volume among participants with detectable calcification (*β*=2.26, p=0.047).

**Conclusion:** This work should provide a practical framework for quantifying ChP calcifications routinely from MRI. The observed relationship between ChP volume and calcification volume highlights the importance of accounting for calcified tissue, particularly when calcification burden is substantial, when investigating ChP abnormalities in aging and neurodegenerative disease.

## Introduction

The choroid plexus (ChP) serves as the most proximal component of the neurofluid circuit, facilitating the filtration of blood plasma through specialized epithelia to secrete cerebrospinal fluid (CSF). Histologically, the ChP consists of a specialized cuboidal epithelial monolayer resting upon a basement membrane, which envelopes a central stroma of loose connective tissue and fenestrated capillaries^1^. With advancing age, the ChP undergoes significant structural remodeling: epithelial cells typically atrophy, while the stroma exhibits progressive collagen deposition, hyalinization, and vascular wall thickening^1–4^. A hallmark of this senescence is the accumulation of laminated calcareous mineralization primarily composed of calcium phosphates and carbonates^5,6^.

ChP is the most common site of physiological intracranial calcification, with prevalence approaching 70% in middle-aged and older adults^7,8^. Because calcified deposits contribute to measured ChP volume and simultaneously compress the choroidal microvasculature on routine neuroimaging sequences, a substantial fraction of the reported hyperplasia and hypoperfusion signal, reported in a wide spectrum of neurodegenerative, neuroinflammatory, vascular, and neuropsychiatric conditions^9–16^, may in fact reflect mineral accumulation rather than functional epithelial remodeling. However, the majority of ChP volumetric and perfusion studies do not attempt to separate calcified from non-calcified tissue, limiting the biological interpretability of these measures and potentially confounding comparisons across cohorts with different calcification burdens. Establishing an MRI-based method to quantify and account for ChP calcification is, therefore, a necessary step for the neurofluid field to move from descriptive associations to mechanistically interpretable biomarkers.

Computer tomography (CT) remains the gold standard for quantifying tissue density, with calcification in the ChP typically exceeding 60 Hounsfield units (HU)^17,18^, but is rarely acquired in MRI-based research and often may not distinguish calcification from blood products, which also appear hyperdense. Quantitative Susceptibility Mapping (QSM) has emerged as an MRI technique that quantifies spatially consistent maps of magnetic susceptibility from local magnetic field variations on appropriately parameterize *R*_2_*-weighted sequences^19,20^. QSM provides a localized, quantitative measure of tissue composition and can differentiate between diamagnetic materials such as calcium, from paramagnetic substances such as iron^21^. QSM offers a non-invasive window into the structural transformation of the ChP.

Although previous studies have used QSM to investigate intracranial calcifications^22,23^, two gaps remain. First, the level of agreement between QSM-derived susceptibility and CT-derived density has not been rigorously quantified within the ChP, limiting how diamagnetic susceptibility should be interpreted as a surrogate for mineral burden. Second, no consensus exists on the susceptibility threshold that reliably separates calcified from non-calcified ChP tissue, or on how such a threshold should correspond to the density cutoffs (60–100 HU) routinely used in CT studies. To address these gaps, we leveraged a cohort of participants who underwent both CT and QSM to evaluate an optimal diamagnetic susceptibility threshold for ChP calcification detection and investigated the extent to which calcification volume accounts for the observed variability in ChP volume.

## Materials and Methods

### Participants

All participants provided informed written consent in accordance with the local institutional review board (IRB) and consistent with the Declaration of Helsinki and its amendments for this prospective study. Inclusion criteria for all participants included no history of cerebrovascular disease (prior stroke or major cervical or intracranial arterial stenosis > 70%), anemia, or psychosis. We deliberately included patients with different ages and meeting clinical criteria for different neurodegenerative conditions to enrich the dataset for calcifications, which are known to increase with age. Clinical assessments were performed by a board-certified neurologist and anatomical imaging and angiography reviewed by a board-certified neuroradiologist.

### Imaging acquisition

All participants were scanned on the same 3 Tesla scanner (Philips Healthcare, Best, The Netherlands) using body coil radiofrequency transmission and phased array 32-channel head coil reception. The protocol included a 3D *T*_1_-weighted magnetization-prepared-rapid-gradient-echo (TR = 8.1 ms, TE = 3.7 ms, spatial resolution = 1.0×1.0×1.0 mm) and a 3D multi-echo gradient echo sequence with flow compensation (TR = 44 ms, first TE = 5.8 ms, echo spacing = 6.7 ms, echoes = 6, flip angle = 18°, spatial resolution = 1.1×1.1×1.1 mm, reconstructed to 0.94 mm isotropic).

CT images were acquired from PET/CT examinations that each participant had undergone as part of a separate PET research protocol ^24,25^. The CT component of these examinations, originally acquired for PET attenuation correction and anatomical localization, was repurposed here for calcification analysis. All PET/CT scans were performed on a Philips Vereos Digital PET/CT scanner (Philips Healthcare, Best, Netherlands), with X-ray exposure ranging from 5 to 30 mSv (FoV = 599.0×599.0×164.0 mm, spatial resolution = 1.2×1.2×2.0 mm).

### Image processing

#### Tissue segmentation

The ChP was automatically segmented from the *T*_1_-weighted images using a previously validated and publicly available deep-learning model^26^. Whole-brain tissue segmentation was performed on the same *T*_1_-weighted image with AssemblyNet^27^.

#### Quantitative susceptibility mapping

Data analyses were performed using MATLAB (Mathworks, Natick, MA), *FSL*, and the Advanced Normalization Tools (ANTs). Phase unwrapping was performed using the Rapid Opensource Minimum spanning trEe algOrithm (ROMEO)^28^. The brain mask was generated automatically from the ROMEO quality map. QSM reconstruction steps were performed using the Morphology Enabled Dipole Inversion (MEDI) toolbox^29,30^. After field map estimation from the unwrapped phase, background field removal was applied using the projection onto dipole fields method^31^, and susceptibility maps were calculated using MEDI with automatic CSF zero referencing^32^. Finally, QSM maps were co-registered to each participant’s T_1_-weighted anatomical scan using ANTs^33^.

#### Computerized tomography

CT images were aligned to *T*_1_-weighted MRI using a two-steps procedure. First, synthesized *T*_1_-weighted MRI were estimated from the CT using SynthSR^34^. Second, the synthetic *T*_1_-weighted images were rigidly registered to the *T*_1_-weighted MRI with ANTs, and the resulting transformation was applied to the original CT. All alignments were visually inspected in ITK-SNAP^35^ to confirm correct co-registration of ChP tissue and calcification between CT and QSM images.

### Quantification of the ChP calcification on CT

Reference calcification within the ChP was defined by combining the ChP mask with the CT image aligned to the *T*_*1*_ ⍰weighted MRI. Using previously reported threshold values, voxels within the ChP mask with HU greater than 60 and 100 were classified as low-density and high-density calcified tissue, respectively, partitioning the ChP into calcified and non-calcified components.

### Estimation of the optimal QSM threshold for calcification detection

The Bayes optimal decision criterion was employed to estimate the optimal susceptibility threshold *τ_χ_*, for detecting mineralization within the ChP at defined density as measure from CT. The threshold was defined as the point where the posterior probabilities of the two tissue classes, calcified (noted *ω_c_*) and non-calcified (noted *ω*_*nc*_), are equal, effectively minimizing the total risk of misclassification. This equilibrium is expressed as:

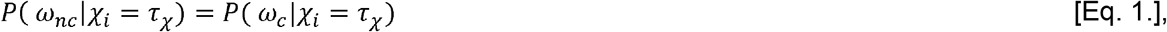

where *χ*_*i*_ represents the observed susceptibility signal from the QSM image at the voxel *i*. Applying Bayes’ theorem, this equality can be rewritten in terms of class-conditional likelihoods and prior probabilities:

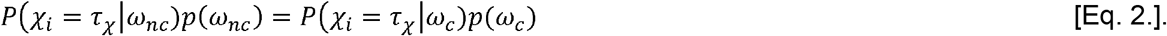

In this work, we assume that the conditional probability *P*(*χ* _*i*_|*ω*_*nc*_) follows either a normal distribution or a Laplacian distribution, while we assume that *P*(*χ* _*i*_|*ω*_*nc*_) follows either a normal distribution or a log-normal distribution. A dual-normal assumption allows for an analytical quadratic solution, however, combinations involving non-Gaussian distributions result in transcendental equations. Consequently, we utilized a numerical optimization approach to find the estimated threshold, noted 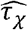 as follows:

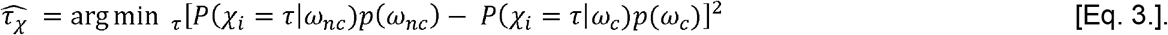

Here, the squared difference serves as the objective function, ensuring a global minimum at the intersection of the two weighted likelihoods. For all experiments, we assumed equal prior weight. Finally, to validate the physical basis of the detected diamagnetic signals, a linear regression was performed to correlate QSM susceptibility values with corresponding HU from CT imaging.

### Implementation and statistics

To evaluate the best-fitted distribution model, the Akaike information criterion (AIC) and Bayesian information criterion (BIC) were calculated between the fitted model and the observed density distribution estimated using the histogram method. Numerical optimization was performed using the Nelder-Mead solver^36^. The optimal threshold was determined within a leave-one-out cross-validation procedure to minimize bias. The resulting thresholds were summarized using the mean and standard deviation.

The calcified ChP volumes obtained from the susceptibility thresholds were compared to those derived from CT images using both 60 and 100 HU thresholds. Agreement was assessed using the intra-class correlation coefficient (ICC; two-way random effects, absolute agreement, single measure) and root mean square error (RMSE).

Finally, we evaluated the relationship between ChP volume and calcification volume to determine how much ChP enlargement can be explained by calcification burden. Mixed-effect linear regression models were used, with ChP volume as the dependent variable, calcification volume as the independent variable, an interaction term for the presence or absence of calcified tissue, intracranial volume (ICV) as a covariate, and subject identifier as the random intercept. Statistical significance was set at p < 0.05.

## Results

### Participants

Twenty adult participants (age=62±12 years) were included, spanning a range of diagnoses: ten with pure autonomic failure, six with Alzheimer disease, three with Parkinson disease, and one asymptomatic healthy control (Table 1). ChP calcifications were quantified on CT using two HU thresholds: 60 HU to capture low-density calcification and 100 HU to capture high-density calcification. Four participants showed no detectable calcification at either threshold, sixteen showed at least one low-density calcification in the left or right ChP, and thirteen showed calcification exceeding 100 HU.

**Table 1.**
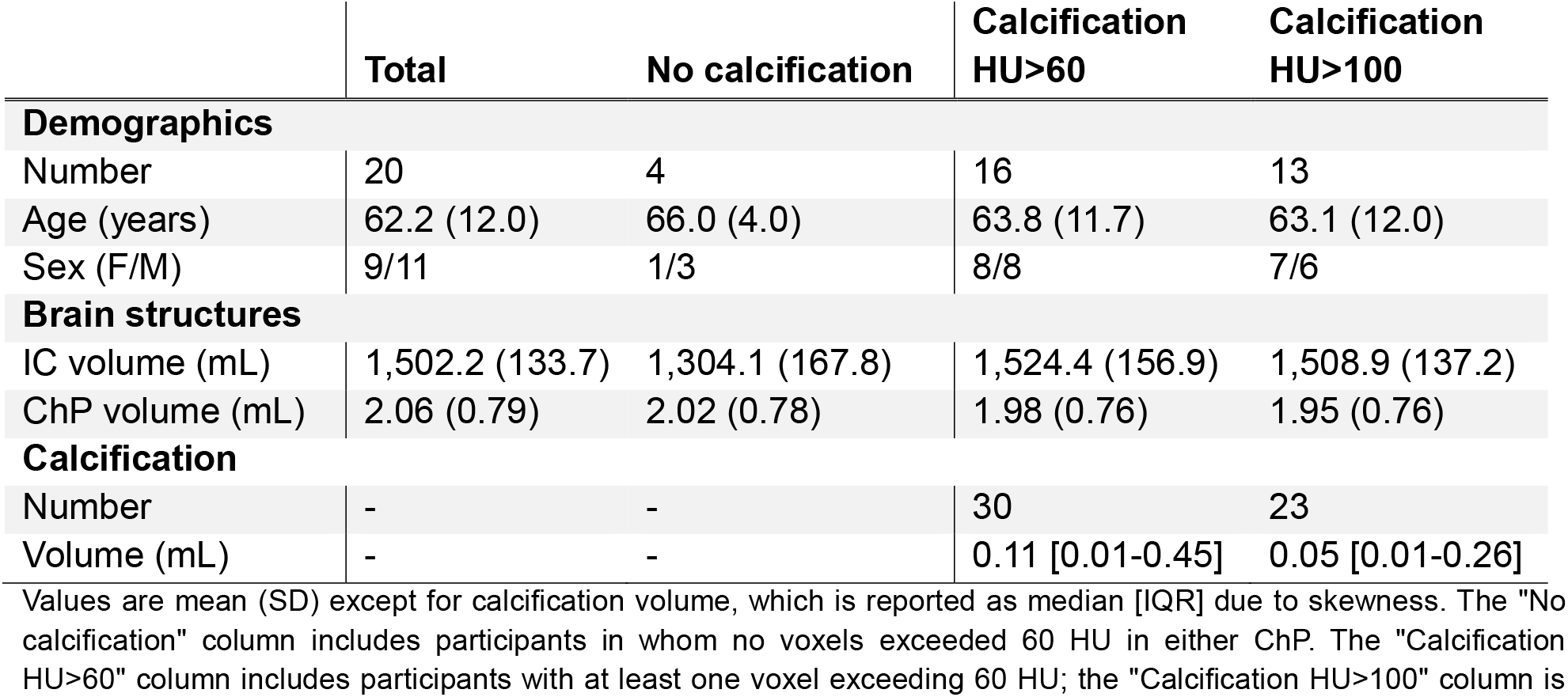
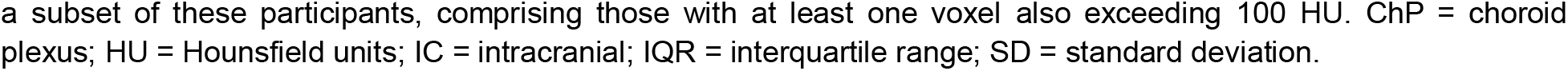
Demographic, brain volumetric, and ChP calcification descriptions.

|  | Total | No calcification | Calcification<br>HU>60 | Calcification<br>HU>100 |
| --- | --- | --- | --- | --- |
| **Demographics** |  |  |  |  |
| Number | 20 | 4 | 16 | 13 |
| Age (years) | 62.2 (12.0) | 66.0 (4.0) | 63.8 (11.7) | 63.1 (12.0) |
| Sex (F/M) | 9/11 | 1/3 | 8/8 | 7/6 |
| **Brain structures** |  |  |  |  |
| IC volume (mL) | 1,502.2 (133.7) | 1,304.1 (167.8) | 1,524.4 (156.9) | 1,508.9 (137.2) |
| ChP volume (mL) | 2.06 (0.79) | 2.02 (0.78) | 1.98 (0.76) | 1.95 (0.76) |
| **Calcification** |  |  |  |  |
| Number | - | - | 30 | 23 |
| Volume (mL) | - | - | 0.11 [0.01-0.45] | 0.05 [0.01-0.26] |

Values are mean (SD) except for calcification volume, which is reported as median [IQR] due to skewness. The "No calcification" column includes participants in whom no voxels exceeded 60 HU in either ChP. The "Calcification HU>60" column includes participants with at least one voxel exceeding 60 HU; the "Calcification HU>100" column is
a subset of these participants, comprising those with at least one voxel also exceeding 100 HU. ChP = choroid plexus; HU = Hounsfield units; IC = intracranial; IQR = interquartile range; SD = standard deviation.

**Table 2.** Statistical summary of the mixed-effect linear model investigating the relationship between ChP volume and calcification volumes in calcified ChP.

|  | Coefficient | SE | z-value | p-value | 95% CI |
| --- | --- | --- | --- | --- | --- |
| **Intercept** | -2.244 | 1.434 | -1.565 | 0.117 | [-5.054, 0.565] |
| **Calcification volume** | 2.258 | 1.136 | 1.987 | 0.047 | [0.030, 4.485] |
| **ICV** | 0.003 | 0.001 | 2.742 | 0.006 | [0.001, 0.005] |
| **RI variance** | 0.287 | 0.669 |  |  |  |

Model fit to hemisphere-level observations from the 16 participants with detectable calcification (32 ChPs). ChP volume (mL) was the dependent variable; calcification volume (mL) was the predictor, with intracranial volume (ICV, mL) as a covariate and participant as a random intercept (Group). Coefficients, standard errors, z- and p-values, and 95% confidence intervals are reported for each fixed effect; for the random effect, the estimated variance is shown. CI = confidence interval; ChP = choroid plexus; ICV = intracranial volume; SE = standard error, RI = random intercept.

### Optimal susceptibility threshold

First, the fit between empirical susceptibility distributions and candidate models was assessed for the Bayes-optimal decision boundary. For non-calcified ChP tissue, the normal distribution yielded AIC = 846.76, BIC = 854.48 (*µ*_*χ*_ = −0.007, *σ*_*χ*_ = 0.032 ppm), while the Laplace distribution gave much lower AIC = 439.79, BIC = 447.51 (*µ*_*χ*_= −0.001 ppm, *σ*_*χ*_ = 0.017 ppm). For calcified tissue, the normal model had AIC = −91.85, BIC = −86.64 (*µ*_*χ*_ = −0.191, *σ*_*χ*_ = 0.142 ppm), but the log-normal model was superior with an AIC = −265.96 and BIC = −260.75 (*µ*_*χ*_ = −0.184, *σ*_*χ*_ = 0.148 ppm). Thus, subsequent analyses used the Laplace distribution for non-calcified and the log-normal for calcified ChP tissue.

Next, we evaluated the agreement of ChP calcification detection using two CT⍰ based density thresholds (Figures 2 and 3): low ✰ density calcification defined by a threshold of 60 HU and high⍰ density calcification defined by a threshold of 100 HU. Agreement metrics were calculated separately for each threshold. The optimal susceptibility threshold was estimated as *τ*_*χ*_ = −0.10 (SD = 0.002) for the 60 HU threshold and *τ*_*χ*_ = −0.15 (SD = 0.003) for the 100 HU threshold.

**Figure 1.**
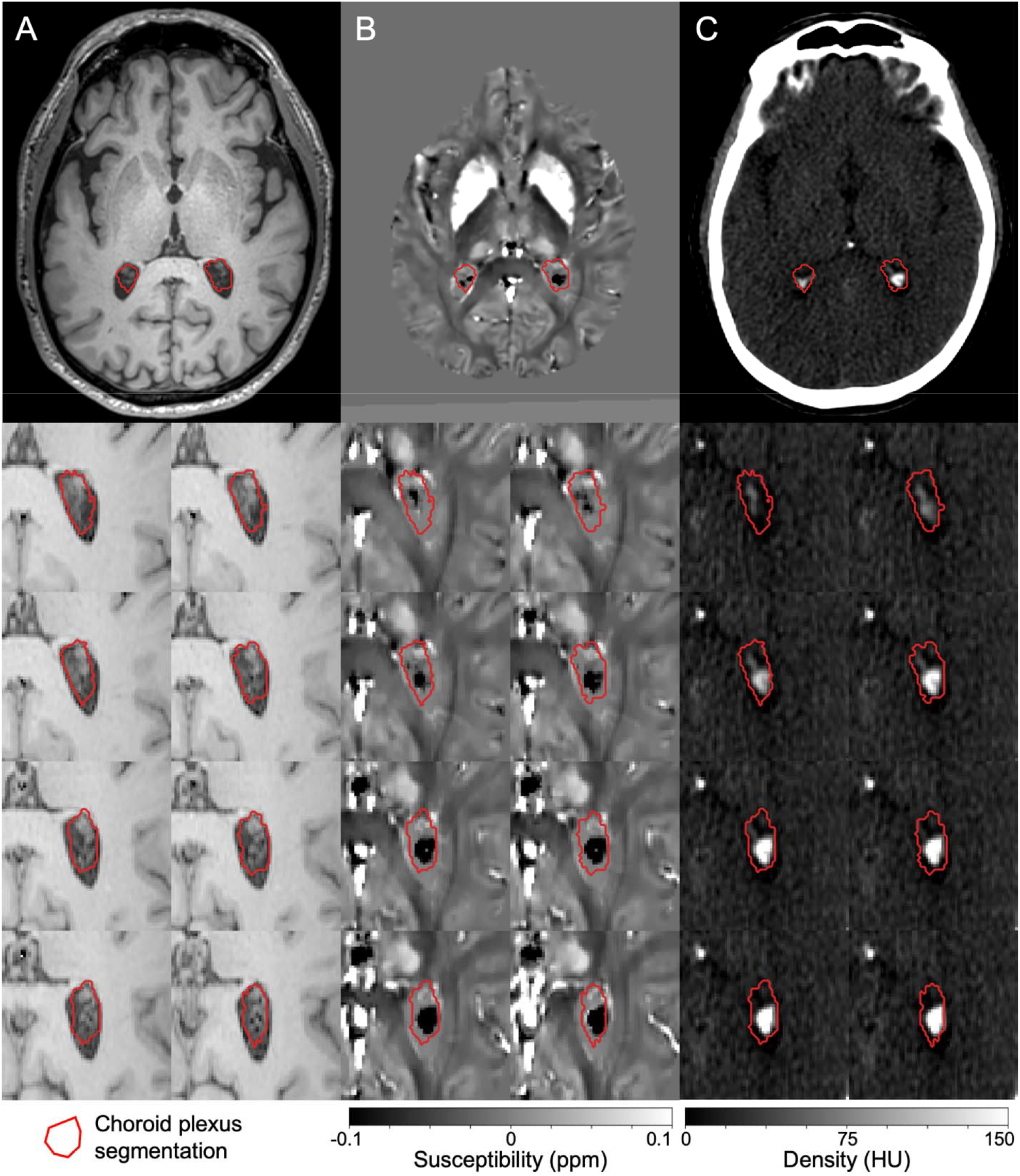
Visualization of choroid plexus calcification. An enlarged choroid plexus is identified on *T*_1_⍰weighted MRI (A), with bilateral calcifications visualized using quantitative susceptibility mapping (QSM; B) and computed tomography (CT; C). Choroid plexus segmentations are overlaid as red contours in each panel. The top row shows full axial views for each imaging modality, while the bottom rows present magnified views of multiple axial slices centered around the left choroid plexus center of mass. Diamagnetic susceptibility appears hypointense on QSM, whereas regions of high density appear hyperintense on CT images.

**Figure 2.**
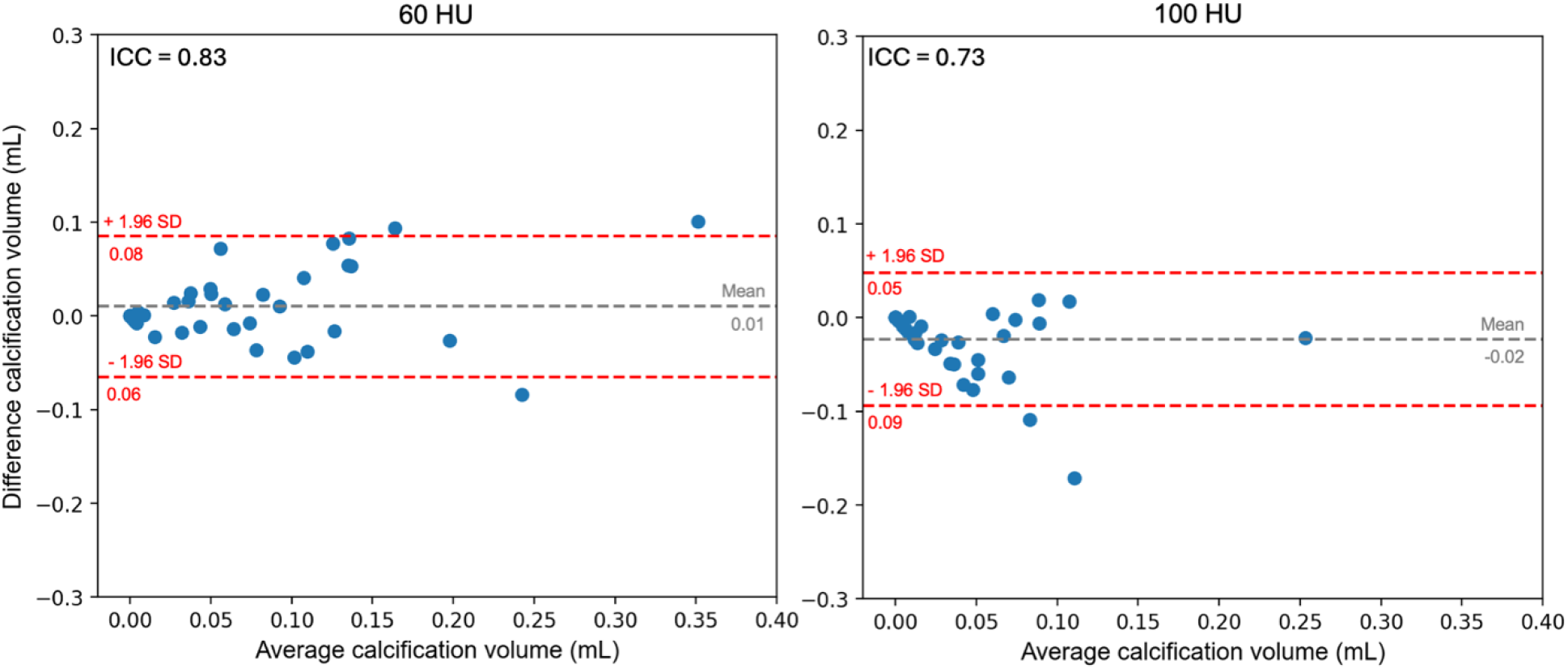
Agreement in calcification detection between modalities. Bland–Altman analysis showing agreement between calcification volumes estimated from CT thresholding and volumes obtained using the estimated optimal threshold. Results are shown for CT density thresholds of 60 HU (left) and 100 HU (right).

**Figure 3.**
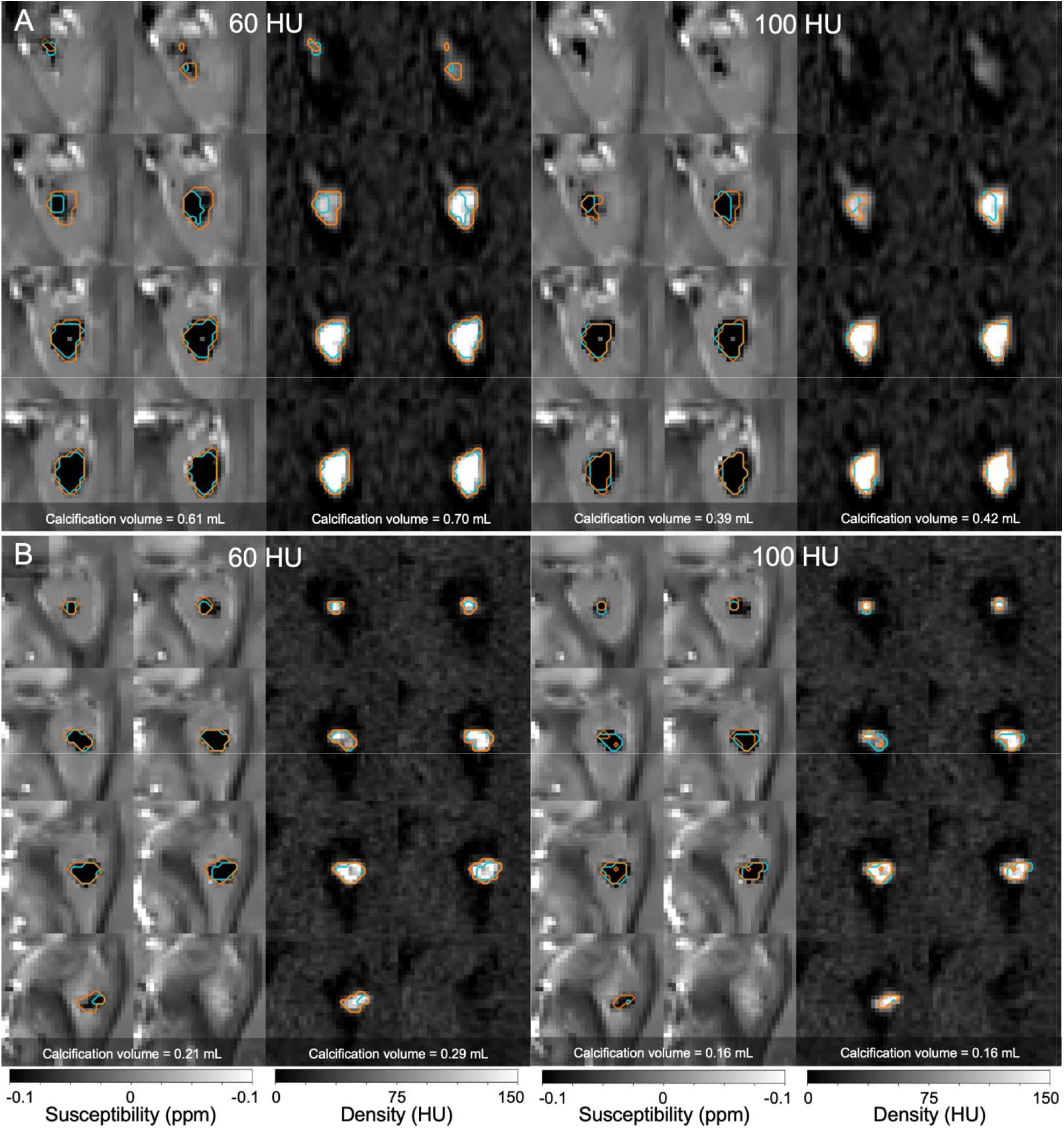
Calcification detection in two participants. Comparison of calcification detection using CT Hounsfield unit (HU) thresholding and a QSM-derived optimal threshold. Segmentation masks are overlaid on zoomed-in axial slices centered on the left choroid plexus, visualized with QSM (left) and CT (right). Panels A show a participant with a large, dense calcification aggregate, whereas panels B show a participant with a medium-sized choroid plexus calcification. Left panels display calcification detected using a density threshold of 60 HU, while right panels show calcification detected using a density threshold of 100 HU. Orange contours represent segmentation maps obtained from CT thresholding, whereas cyan contours represent segmentation maps obtained using the QSM optimal threshold.

At the low-density threshold of 60 HU, calcification volume measured from CT averaged 0.8 mL (SD = 0.9), compared with 0.7 mL (SD = 0.8) estimated from QSM, with no significant difference between modalities (p = 0.44). Inter⍰ modality agreement was high, with an ICC of 0.83 (p < 0.001) and a root mean square error (RMSE) of 0.03 mL (SD = 0.04).

At the high-density threshold of 100 HU, the average calcification volume measured from CT was 0.030 mL (SD = 0.05), compared with an average of 0.036 mL (SD = 0.05) estimated from QSM. Inter ⍰ modality agreement remained high to moderate, with an ICC of 0.73 (p < 0.001) and an RMSE of 0.02 mL (SD = 0.03).

### Relationship between HU and susceptibility values within the calcified ChP tissue

We investigated the relationship between density measured on the low-dose CT and susceptibility measured on the QSM at the voxel level. Using a linear mixed-effect model, where diamagnetic susceptibility was set as the dependent variable, density value as the independent variable, and random intercept set as the image identification, we observed a significant relationship between the diamagnetic signal and density (*β*=-0.0026, standard error= 9.26 10^-05^, p<0.01).

### Choroid plexus volume and calcification

Using a mixed⍰effects linear regression model, we observed a positive association between ChP volume and calcification volume in ChPs exhibiting calcification at the low⍰density threshold (*β* = 2.26, standard error = 1.14, p = 0.047). However, ChP volume did not differ between the four participants without detectable calcification and the 16 participants with calcification (mean difference: 0.04 mL, p = 0.88). Thus, although calcification burden scales with ChP volume within calcified participants, ChP volume alone does not distinguish participants with and without detectable calcification.

## Discussion

The ChP has recently gained substantial interest due to its important roles in neuroimmune functions and CSF production, with most studies focused on investigating volume and perfusion changes. However, the presence and prevalence of calcification might have a fundamental impact on how volume increases and perfusion decreases are interpreted. Here, we used QSM to detect calcifications within the ChP tissue and provided interpretable susceptibility thresholds that link QSM to the CT tissue-density measures classically used to characterize ChP calcification, using a rigorous statistical modelling framework.

### Methodological evaluation and integration with previous literature

Choroid plexus calcifications are typically detected post-mortem by microscopy or in vivo using CT, which provides quantitative and interpretable tissue density measures. Studies have used CT thresholds ranging from 60 to 100 HU to assess calcification^17,18^. In this work, we evaluated the relationship between CT-derived density and QSM-derived susceptibility within the ChP, finding a linear correlation between these modalities. The susceptibility values measured in calcified ChP were consistent with previous whole-brain calcification studies^22,23^.

To determine the optimal susceptibility threshold, we adopted a Bayes⍰ optimal decision framework. The validity of this approach relies on the correct specification of the class⍰conditional probability density functions and class priors. To ensure appropriate model specification, we evaluated several candidate distribution models for each tissue class and selected the distributions that best fit the empirically observed susceptibility data before estimating the Bayes⍰ optimal threshold. Our experiments showed that the observed susceptibility signal follows a Laplace distribution centered at zero, reflecting the lack of net paramagnetic or diamagnetic material in non-calcified ChP and the presence of sparse, heavy-tailed variability from susceptibility reconstruction and anatomical heterogeneity. Unlike the normal model, the Laplace distribution better captures the sharp central tendency and high kurtosis seen empirically, providing a more robust description of background fluctuations. For calcified ChP tissue, susceptibility values were best fit by a log-normal distribution, consistent with the heterogeneous and progressive nature of mineral deposition. This distribution reflects the multiplicative effects of calcium burden, partial-volume, and microstructural variation, resulting in asymmetric, heavy-tailed susceptibility values compared to the symmetric, noise-dominated fluctuations in non-calcified tissue.

Finally, within the detected ChP calcification mask, susceptibility values were consistent with previous reports. For instance, a previous study reported a mean susceptibility of −0.21□ppm, which is slightly more negative than the susceptibility estimated using the normal distribution model in this study (i.e., −0.18□ppm)^23^. It is noteworthy that mean values reported by previous studies assume symmetry of the susceptibility distribution, an assumption that was not supported by our data. In contrast, the log⍰ normal distribution modeling yielded to values aligned with the median value previously reported (i.e., −0.19□ppm) without assuming distributional symmetry. This further supports the generalizability of the proposed susceptibility threshold and its potential utility in future studies aimed at quantifying the progression of ChP calcification volume and interpreting these changes in the context of other physiological measures.

### Clinical relevance and implication in current methodology

The high prevalence of calcifications raises a volume-function paradox that complicates MRI interpretation. These non-functional deposits might partially contribute to the apparent volumetric enlargement through accumulation of metabolic debris and fibrosis rather than an increase in secretory epithelial cells. Simultaneously, these dense structures could physically compress the choroidal microvasculature, directly driving the perfusion decreases seen in ASL imaging. Consequently, what might appear as enlargement on standard MRI could be a marker of degenerative remodeling, in which the ChP is physically larger but functionally failing due to structural overcrowding and reduced vascular access.

Importantly, recent work has demonstrated that deep learning algorithms can be applied to routine, non-contrast brain imaging scans^26,37^. This work expands on this segmentation infrastructure to demonstrate these regions can additionally be decomposed into calcified and non-calcified regions with the addition of a QSM protocol and the proposed analysis pipeline.

### Limitations

Accurate co-registration of CT images to T1-weighted MRI is essential for valid voxel-to-voxel comparisons between QSM and CT signals. To optimize alignment, we used a specialized pipeline that synthesized T1-weighted MRI from CT data, enhancing inter-modality similarity. Images were visually inspected, and those not meeting strict alignment criteria were excluded. While CT is the standard for calcification detection, it cannot reliably distinguish mineralization from blood products, as both show high attenuation. To address this, we used QSM to identify paramagnetic hemorrhages and excluded participants with paramagnetic ChP signals, ensuring masks captured only diamagnetic calcified tissue.

## Conclusions

We established a principled MRI⍰based framework for detecting ChP calcification by defining optimal diamagnetic susceptibility thresholds. By explicitly modeling class⍰ conditional susceptibility distributions for non-calcified and calcified ChP tissue, we derived susceptibility cutoffs that minimize expected misclassification risk and demonstrate strong agreement with CT⍰based density measures across both low⍰ and high⍰ density calcification regimes. This approach enables calcification detection using QSM to move beyond empirically chosen or scanner⍰ dependent thresholds toward a statistically grounded and reproducible framework that can be validated and refined in larger, multi-site cohorts.

## Data Availability

All data produced in the present study are available upon reasonable request to the authors.

